# Impact of complete lock-down on total infection and death rates: A hierarchical cluster analysis

**DOI:** 10.1101/2020.05.03.20089649

**Authors:** Samit Ghosal, Rahul Bhattacharyya, Milan Majumder

**Author notes:** **Corresponding author:** Samit Ghosal.

## Abstract

**Introduction and Aims:** Retarding the spread of SARS-CoV-2 infection by preventive strategies is the first line of management. Several countries have declared a stringent lock-down in order to enforce social distancing and prevent the spread of infection. This analysis was conducted in an attempt to understand the impact of lock-down on infection and death rates over a period of time.

**Material and Methods:** A validated database was used to generate data related to countries with declared lock-down. Simple regression analysis was conducted to assess the rate of change in infection and death rates. Subsequently, a k-means and hierarchical cluster analysis was done to identify the countries that performed similarly. Sweden and South Korea were included as counties without lock-down in a second-phase cluster analysis.

**Results:** There was a significant 61% and 43% reduction in infection rates 1-week post lock-down in the overall and India cohorts, respectively, supporting its effectiveness. Countries with higher baseline infections and deaths fared poorly compared to those who declared lock-down early on. Sweden and South Korea fared equally well, as most lock-down countries stemmed the growth of infection and death.

**Conclusion:** Lock-down has proven to be an effective strategy is slowing down the SARS-CoV-2 disease progression exponentially. However, lessons need to be learned from Sweden and South Korea on arresting the disease progression without imposing such stringent measures.

## 1.0 Introduction

The SARS-CoV-2 pandemic has raged throughout the world, with all its fury. The scientific and political communities have reacted to this challenge with a varied degree of urgency. The initial days were mired by a lack of accurate and adequate recommendations. This was related to the fact that this was a novel virus and nothing was known about it. However, with the scientific community reacting aggressively to the situation, we got a fairly good understanding of how the virus attaches to the host cells, causes organ damage, and probable treatment targets. This resulted in a flurry of studies conducted in order to arm the healthcare sector. We saw Chloroquine, hydroxychloroquine, and resmdesivir, being extensively tested as possible treatment strategies.[1,2,3] The trials for an effective vaccine are also ongoing. [4] However, in the interim period we need strategies in place to prevent the spread of infection. Social distancing and hygiene became the targets for preventive strategies. Several countries, including India, went ahead with a stringent lock-down.[5]

There is absence of any prior data on the impact of lock-down on prevention of infection. It is of paramount importance to assess the effectiveness of this strategy as the lock-down continues. This study was conducted to analyze the benefits of lock-down on total infection and death rates. Such data are required urgently to aid policymakers into taking a decision on future strategies.

## 2.0 Materials & Methods

Data on infection and death rates were collected from the Worldometer website [6] Details related to the lock-down declaration in different countries were acquired from online media resources [5] The collected data were transferred to a comma separated value (CSV) file, which was then uploaded to Jupyter notebook and analyzed with Python 3.8.2 software (Windows 10 64 bit, USA). A total of 12 countries, including India were taken up for analysis. (Table1) China was left out in view of the restructuring of data. A few other countries also declared lock-down; however, in view of a lot of missing data, they were excluded from the analysis. We intended to include a parallel comparative arm (countries without complete lock-down). However, due to a variable definition of restrictions imposed in most of the other countries, we could include only Sweden and South Korea (countries with minimal degrees of restrictions) as the additional arm. Instead of using both these countries as comparators, we decided to include them in a cluster analysis and visualize where exactly they fit in.

**Table 1:** Countries and dates when the initial lock-down was declared. Baseline characteristics

| Date of lock-down | Initial Lock-down Duration Declared (Days) | Country | Baseline Total infection | Baseline death | Baseline testing frequency/million |
| --- | --- | --- | --- | --- | --- |
| 25th March | 21+ | India | 657 | 12 | 482 |
| 9th March | 21+ | Italy | 9172 | 463 | 29071 |
| 23rd March | 25+ | UK | 6650 | 335 | 9867 |
| 14th March | 40+ | Spain | 6391 | 196 | 28779 |
| 17th March | 14+ | France | 7730 | 175 | 7103 |
| 20th March | 9+ | Germany | 19848 | 68 | 24738 |
| 16th March | 26+ | Austria | 1018 | 3 | 25819 |
| 17th March | 30+ | Belgium | 1243 | 10 | 18468 |
| 11th March | 14+ | Hungary | 13 | 0 | 6793 |
| 13th March | 30+ | Poland | 68 | 2 | 7870 |
| 16th March | 28+ | Malaysia | 566 | 0 | 4291 |
| 25th March | 30+ | New Zealand | 283 | 0 | 25698 |

| **Date of lockdown** | **Initial Lockdown Duration Declared (Days)** | **Country** | **Baseline Total infection** | **Baseline death** | **Baseline testing frequency/million** |
| --- | --- | --- | --- | --- | --- |
| 25th March | 21+ | India | 657 | 12 | 482 |
| 9th March | 21+ | Italy | 9172 | 463 | 29071 |
| 23rd March | 25+ | UK | 6650 | 335 | 9867 |
| 14th March | 40+ | Spain | 6391 | 196 | 28779 |
| 17th March | 14+ | France | 7730 | 175 | 7103 |
| 20th March | 9+ | Germany | 19848 | 68 | 24738 |
| 16th March | 26+ | Austria | 1018 | 3 | 25819 |
| 17th March | 30+ | Belgium | 1243 | 10 | 18468 |
| 11th March | 14+ | Hungary | 13 | 0 | 6793 |
| 13th March | 30+ | Poland | 68 | 2 | 7870 |
| 16th March | 28+ | Malaysia | 566 | 0 | 4291 |
| 25th March | 30+ | New Zealand | 283 | 0 | 25698 |

Inputs: Total infection and death rates at baseline and weeks prior to the lock-down.

Outputs: Rates of total infection and death change at the end of 4 weeks of the lock-down period.

## 3.0 Results

### 3.1 Pre-cluster analysis

#### 3.1.1 Overall regression analysis (lock-down countries only)

Prior to performing the cluster analysis, we assessed the weekly change in infection and death rates using the equation (x-y)/y, where x represents the infection or death at week n and y represents the infection or death at week n-1. Having determined all the rates of change over the 4 weeks period on a weekly basis, a simple regression analysis was done to determine the direction and strength of the change. There was a very strong exponential decay in both the rates of infection (R^2^: 0.995) and death (R^2^: 0.979) over time after lock-down was declared in the overall cohort. (Figure 1) Comparing infection rates a week before and a week after lock-down, resulted in a 61% decrease in infection rate. In India, there was a 45% exponential reduction in rates of SARS-CoV-2 infection rates, indicating a clear benefit of lock-down.

**Figure 1:**
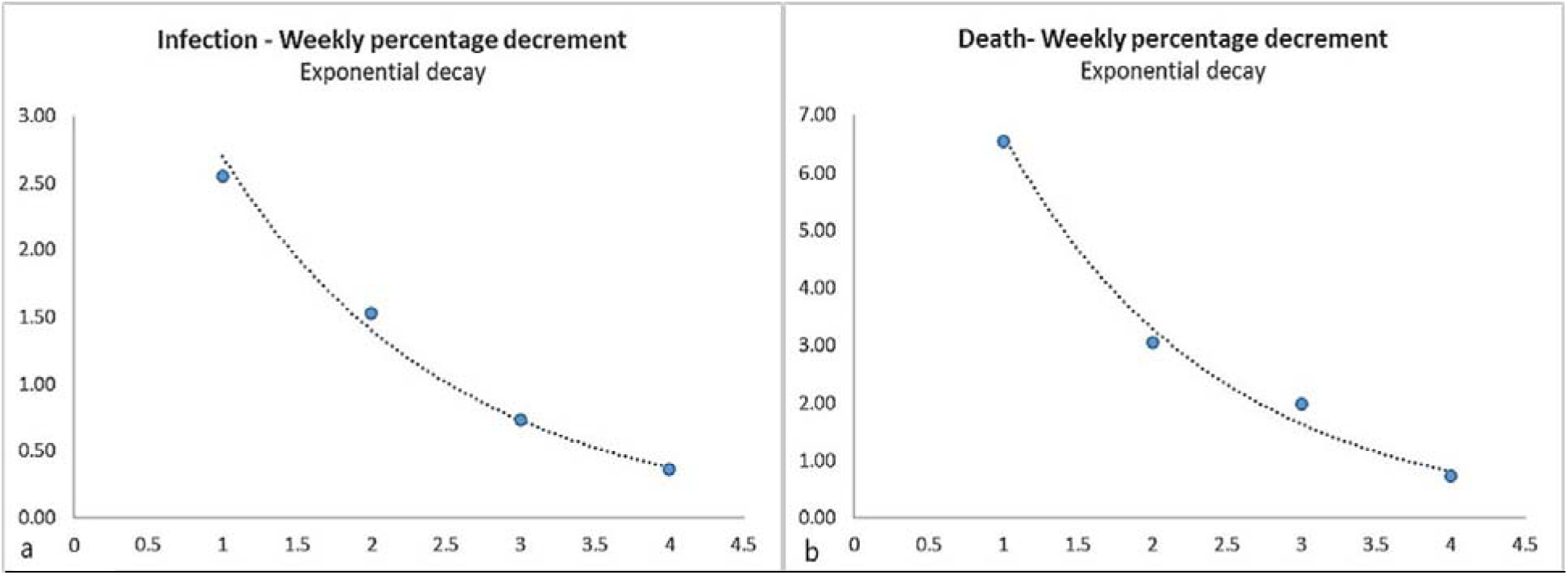
Regression analysis of infection (a) and death (b) rates over a period of 4 weeks. Y-axis: rate of change; x-axis: weeks. Both indices demonstrated an exponential decay. The model for infection rate is summarized by y=5.1988e^−0.655x^ and for death rate by y=13.357e^−0.7x^.

#### 3.1.2 Individual country-wise regression analysis (lock-down countries only)

The strength of exponential decay for infection was stronger than that of death rates. This was evident on visualizing the regression graph for each country individually. (Figure 2 a & b) Unlike reduction in infection rates, reduction in death rates indicated a random pattern of decay for some countries (Figure 2 a).

**Figure 2:**
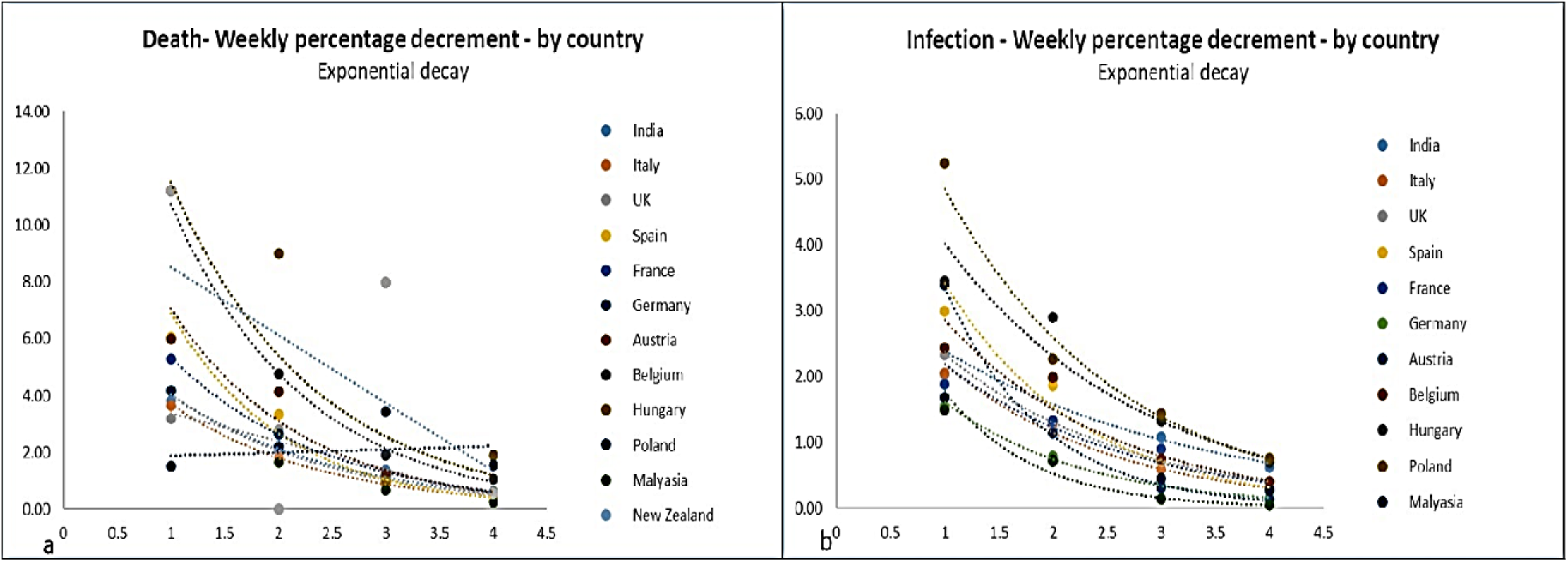
Regression analysis of death (a) and infection (b) rates over a period of 4 weeks-country-wise axis: rate of change; x-axis: weeks. Some countries have random pattern of decay (fig 2 b).

### 3.2 Hierarchical cluster analysis (Lock-down countries)

In view of the differences in the rate of decrease of deaths in some countries, we performed a hierarchical cluster analysis to identify countries with similar rates of decay, taking into account all the input variables. A k-mean hierarchical clustering strategy was employed to identify the appropriate number of possible clusters with minimal variability. The optimum number of clusters identified for analysis was 2, which was decided by the elbow method. (Figure 3)

**Figure 3:**
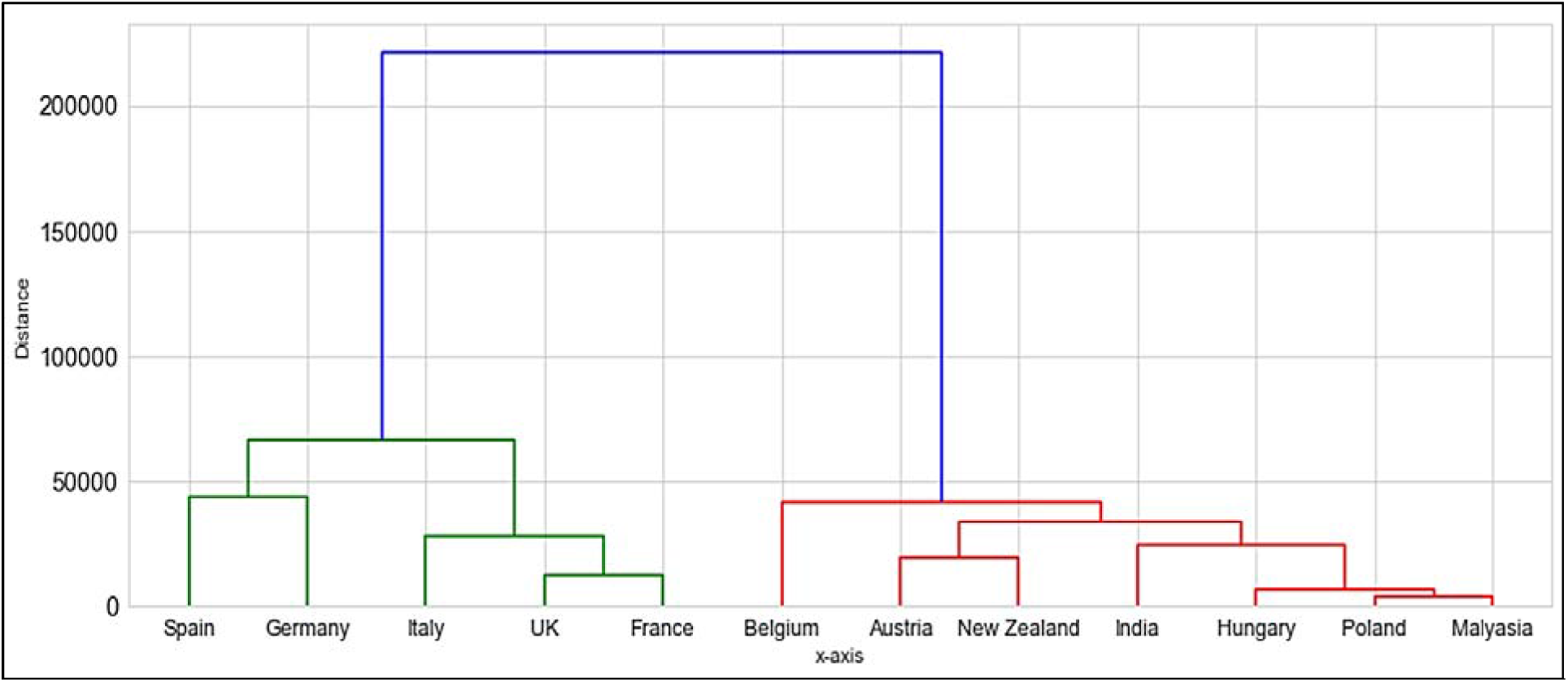
Hierarchical clustering of death rates of the 15 countries with lock-down divided into two clusters. X-axis: countries; y-axis: distance from the mean. Cluster 1 (Green): Spain, Germany, Italy, UK, and France. Cluster 2 (Red): Belgium, Austria, New Zealand, India, Hungary, Poland and Malaysia.

Spain, Germany, UK, France, and Italy were clustered into one group (Cluster 1), whereas Austria, New Zealand, Poland, Malaysia, Hungary, India, and Belgium were clustered into a separate group (Cluster 2). Considering the distance from the mean, cluster 2 benefited more from cluster 1 from the lock-down. Within cluster 2 countries like Malaysia, Poland, and Hungary fared the best, followed by India, Austria, and New Zealand.

Having identified the two separate clusters, we performed the regression analysis on infection and death rates separately on clusters 1 and 2. Cluster 1 demonstrated a very similar and strong trend towards exponential decay in infection (R^2^: 0.989) and death rates (R^2^: 0.987).

However, in cluster 2, there was a slight discordance between the exponential decay in infection rate (R^2^: 0.0.995) and death rate (R^2^: 0.964). This discordance was driven by an increase in death rates from week 2 to week 3 post lock-down in New Zealand.

The decrease in infection rate looked more impressive in cluster 2.

### 3.3 Hierarchical cluster analysis (Lock-down countries plus Sweden & South Korea)

In view of the non-availability of comparative indices (countries without lock-down) due to extreme variations in the definition of non-lock-down, we included two countries (Sweden and South Korea) with minimal restrictions in place in the new cluster analysis. The new hierarchical cluster analysis resulted in Sweden and South Korea falling in cluster 2 as far as decay in infection and death rates were concerned. Within the new cluster 2, the highest reduction in infection and death rates occurred in Malaysia, Poland, and Hungary (lock-down countries), followed by the non-lock-down countries (Sweden and South Korea). (Figure 4) India, Austria, Belgium, and New Zealand fared better than cluster 1, but the rates of death and infection were higher than those of non-lock-down countries and Malaysia, Poland, and Hungary (lock-down countries from cluster 2).

**Figure 4:**
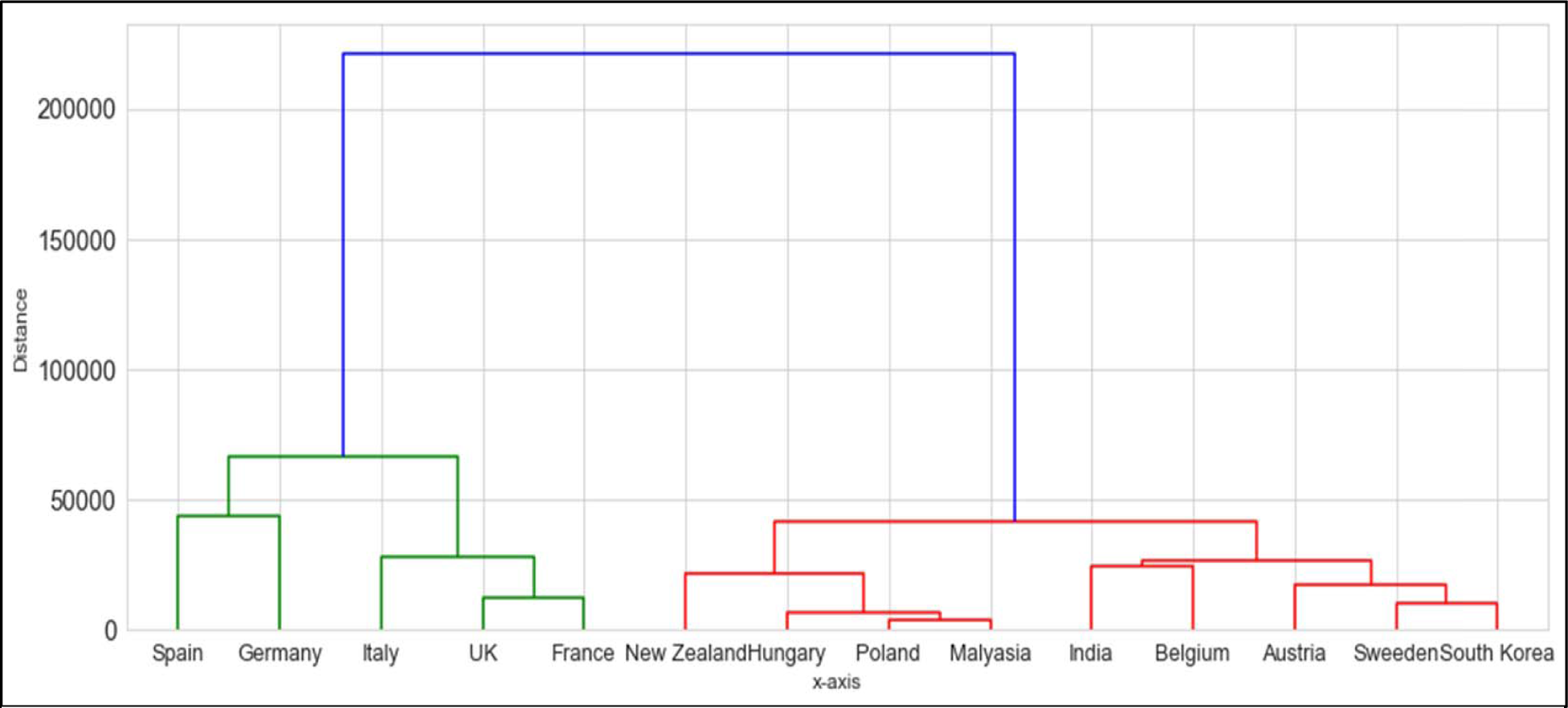
Hierarchical clustering of death rates of the 17 countries (15 with lock-down & 2 without lock down divided into two clusters). X-axis: countries; Y-axis: distance from the mean. Cluster 1 (Green): Sp Germany, Italy, UK, and France. Cluster 2 (Red): Belgium, Austria, New Zealand, India, Hungary, Poland, Malaysia, Sweden and South Korea.

## 4.0 Discussion

### 4.1 What we already know

The SARS-CoV-2 pandemic has proven to be an immense challenge for humanity. In the absence of any definitive treatment modalities and a vaccine, the only effective strategy at present seems to be preventive in nature. A variety of preventive strategies have been put in place by different governments across the world. Some have increased the frequency of testing in an attempt to identify asymptomatic carriers, whereas others have gone for symptom-based screening. [7] Home isolation, quarantine strategies, and complete lock-down are other social distancing strategies employed. Complete lock-down has been the toughest social isolation measure taken to date. It is projected to be the most effective measure preventing the spread of infection, whereas there are significant economic and social downsides associated with it.[8] Lock-down has been declared to be working well by different governments across the world, resulting in extension of the lock-down periods. Some countries, including India, have gone for phased relaxation of lock-down by identifying several districts as hotspots.

### 4.2 What this study adds to the existing information

The claims related to success of the lock-down are presented as the rate of doubling of infection over a period of time. We performed a more objective analysis looking into the impact of lock-down on the progression of total infection and death on a weekly basis. We identified 12 countries where lock-down was declared and also had data for analysis for 4 weeks prior to and after the lock-down date.

Initial simple regression analysis revealed a significant 61% reduction in total infection a week after the lock-down. For India, the reduction of total infection was 45% at week 1 post lock-down. The 4 weeks post lock-down analysis revealed an exponential and significant (R^2^:0.995) decrease in total infection across all the countries who declared a lock-down. The trend was a bit different in the case of deaths with a few countries not showing an exponential decrease. This was in view of the fact that countries like New Zealand show a trend towards an increase in death rates between week 2 and 3 post lock-down.

In order to get a clear picture of how a group of countries behaved, we performed a k-means and a hierarchical cluster analysis. We could club similarly behaved counties into two prominent clusters. Cluster 1 included Spain, Germany, Italy, UK, and France, whereas cluster 2 included Belgium, Austria, New Zealand, India, Hungary, Poland, and Malaysia. Among the two clusters, cluster 1 fared better than cluster 2 as far as overall performance (decreasing infection and death rates) was concerned. This could be explained by the higher total infection and death counts in the cluster 2 countries at baseline (lock-down date). (Table 1)

We performed a repeat analysis of hierarchical clustering, including Sweden and South Korea as countries with minimal restrictive measures in place. Since the inclusion of just 2 countries as direct comparative arms would have generated skewed data, we included these countries as a part of the pooled analysis with the other 12 countries. The total infection and death rate also decayed exponentially in these countries, comparable to cluster 2. Malaysia, Poland, and Hungary demonstrated the best response, followed by the other countries in cluster 2, including Sweden and South Korea. This difference could also be explained by the lower baseline total infection and death in Malaysia, Poland, and Hungary compared to Sweden and South Korea.

The similarity of response between Sweden and South Korea and India, Austria, Belgium, and New Zealand could prompt us into question the effectiveness of lock-down in later countries. The two non-lock-down countries started with higher baseline total infections and deaths than India. The exponential decay in both these countries could be attributed to effective government policies and better implementation of social distancing as well as testing strategies.

### 4.3 Limitations of this study

Our study was limited by the non-inclusion of several other countries with lock-down, which could have steered the results in a different direction. The second limitation is related to the absence of a direct comparative arm. This could have given a better understanding of the direction of this pandemic. Third, apart from total infection, deaths, and testing frequency, there are other variables both objective as well as subjective, which could have influenced the outcomes.

### 4.4 Strengths of the study

This is the first in a kind analysis looking into the pattern of change in the two most important parameters followed in this pandemic. Although a direct comparative arm would have been of great value, the lack of a clear-cut definition of partial lock-down prompted us to leave these countries out of our analysis. Although other variables could have influenced the results differentially, the regression models were very robust, supporting the included inputs.

## 5.0 Conclusion

This is the first study to highlight the impact of lock-down on infection and death rates over a period of 4 weeks. There was an exponential decrease in both infections as well as death with lock-down.

The declaration of lock-down early on in the pandemic proved to be a more effective measure. However, two countries (Sweden and South Korea) performed equally well. This could be an opportunity to assess the policies in place in Sweden and South Korea, implement them, and come out of the lock-down.

## Data Availability

All data are freely available online.

